# Using Real-Time Machine Learning to Prevent In-Hospital Severe Hypoglycemia: A prospective study

**DOI:** 10.1101/2022.07.21.22277774

**Authors:** Michael Fralick, Meggie Debnath, Chloe Pou-Prom, Patrick O’Brien, Bruce A. Perkins, Esmerelda Carson, Fatima Khemani, Muhammad Mamdani

**Affiliations:** Sinai Health System, Division of General Internal Medicine, Toronto, Ontario; Data Science and Advanced Analytics, Unity Health, Toronto, Ontario; Sinai Health System, Division of Endocrinology, Toronto, Ontario; Institute of Health Policy, Management and Evaluation, University of Toronto, Toronto, Ontario, Canada; St Michael’s Hospital, Division of Vascular and Cardiovascular Surgery, Unity Health, Toronto, Ontario

## Abstract

**Objective:** There are many examples of machine learning based algorithms with impressive diagnostic characteristics. However, a few published studies have evaluated how well they perform when deployed into clinical care. The objective of this study was to evaluate the performance of a recently validated machine-learned model to predict inpatient hypoglycemia following its implementation into clinical care on cardiovascular and vascular surgery ward.

**Methods:** We conducted a prospective analysis of a machine learning algorithm to predict hypoglycemia. The algorithm was trained, validated, and tested using data from 2013 to 2019. We employed multiple supervised machine learning techniques (e.g., extreme gradient boosting) to predict inpatient hypoglycemia and severe hypoglycemia using a wide-range of patient-level data (i.e., features) including medications, labs, nursing notes, comorbid conditions, among others.

**Results:** Our study included 3989 hospitalizations during the pre-implementation period and 1916 post-implementation. Approximately one-third of patients were women, the median age was 66 years, 23% received metformin in hospital, 7% received a sulfonylurea, and the median length of stay was 6 days. During the pre-implementation period, more than 5% of patients experienced hypoglycemia during 9.4% (N=12/127 weeks) of study weeks as compared to 0% (N=0/79 weeks) of weeks during the post-implementation period (p=0.012). The weekly variability in the rates of hypoglycemia decreased by approximately 50% from the pre-implementation (standard deviation 1.8, variance 3.4) to implementation phase (standard deviation 1.3, variance 1.6; p=0.03). There was a week-to-week decrease in hypoglycemia rates by 0.03 events per week [95% CI: -0.04, -0.01] (p = 0.004) but no significant change in weekly rates of hyperglycemia (−0.04 [95% CI: -0.10, 0.01]; p=0.102). The severe hypoglycemia events per 100 patients per year was 1.3 pre-implementation and 1.1 following implementation.

**Discussion and Conclusion:** Our prospective analysis of a recently validated machine learned model to prevent hypoglycemia demonstrated a reduction in the rates of inpatient hypoglycemia. Our study suggests that machine learning methods can be leveraged to prevent inpatient hypoglycemia.

## INTRODUCTION

There are many examples of machine learning based algorithms with impressive diagnostic characteristics,^1^ but few published studies have evaluated how well they perform when deployed into clinical care.^2^ We recently published the results of a machine-learned model developed to predict inpatient hypoglycemia.^3^ The objective of our current study was to evaluate its performance following implementation into clinical care on cardiovascular and vascular surgery ward. Patients on these wards are at particularly high risk of hypoglycemia because guidelines recommend tight glycemic control post-operatively^4^.

## METHODS

We conducted a prospective analysis of a machine learning algorithm to predict hypoglycemia. The algorithm was trained, validated, and tested using data from 2013 to 2019. The details of the machine learning methods have been published, but in brief we employed multiple supervised machine learning techniques (e.g., extreme gradient boosting) to predict inpatient hypoglycemia and severe hypoglycemia using a wide-range of patient-level data (i.e., features) including medications, labs, nursing notes, comorbid conditions, among others. Our deployed model was an extreme gradient boosting model.^3^

The pre-implementation period for the model was Jan 1, 2018 to May 31, 2020 and the model was implemented on the cardiovascular surgery and vascular surgery ward at St. Michael’s Hospital of Unity Health January 1, 2021 and evaluated until April 30, 2022. Over 50% of patients on these wards have diabetes. Prior to implementation we met with the nurse practitioners to understand how best to provide them with the results of the algorithm. The nurse practitioners are responsible for the day-to-day clinical care of patients during their hospitalization from Monday to Friday. They requested that a daily email that included the names of the patients at highest risk of hypoglycemia would be the ideal approach. The email itself was generated by the algorithm and thus was entirely automated. The email includes a list of patients identified as highest-risk by the model and an additional list of patients that experienced a blood glucose level below 6.0 mmol/L in the previous 24 hours. This was to support the nurse practitioners in reviewing both patients anticipated for a hypoglycemia event and those who were actively trending low independent of the algorithm. The intent was to support our clinicians with daily actionable information for patients that were identified as high-risk at that point in time. No other information was provided in the email such as approaches to reduce the risk of hypoglycemia. This was purposeful because our end-users were already experts in preventing hypoglycemia based on their years of clinical experience.

Our primary outcome was the proportion of weeks before and after model implementation where more than 5% of patients experienced hypoglycemia (glucose < 3.9 mmol/L) per week. Given the small number of patients on the study units we anticipated significant variability in rates of hypoglycemia and aggregated to weekly estimates. A clinically relevant metric of proportion of patients on the units experiencing hypoglycemia was chosen. The rates were calculated as the sum of (# encounters experiencing hypoglycemia, each day of the week) divided by sum of (daily patient census, for each day of the week) * 100. We used daily event and patient census estimates since the intervention was delivered on a daily basis. We assessed changes in the primary outcome graphically and using a Chi square test. Secondary outcomes included changes in the variance of hypoglycemia rates, weekly rates of hypoglycemia annual rates of severe hypoglycemia (glucose < 2.2 mmol/L), and weekly rates of hyperglycemia. Because severe hypoglycemia is rare we assessed the rate on a yearly basis. Levene’s test was used to assess changes in variance between the pre-implementation and post-implementation periods and segmented regression was used to examine changes in weekly rates of hypoglycemia and hyperglycemia^5^. All statistical analyses were performed using R version 3.6.3.

## RESULTS

Our study included 3989 hospitalizations during the pre-implementation period and 1916 post-implementation. Baseline characteristics of patients, including comorbid conditions, were similar prior to and following implementation (Table 1). Approximately one-third of patients were women, the median age was 66 years, 23% received metformin in hospital, 7% received a sulfonylurea, and the median length of stay was 6 days. In Figure 1, we provided a visual representation of the changes in the rate of the outcomes overtime. Following implementation of our model, we observed reductions in the rate of hypoglycemia. During the pre-implementation period, more than 5% of patients experienced hypoglycemia during 9.4% (N=12/127 weeks) of study weeks as compared to 0% (N=0/79 weeks) of weeks during the post-implementation period (p=0.012). The weekly variability in the rates of hypoglycemia decreased by approximately 50% from the pre-implementation (standard deviation 1.8, variance 3.4) to implementation phase (standard deviation 1.3, variance 1.6; p=0.03). There was a week-to-week decrease in hypoglycemia rates by 0.03 events per week [95% CI: -0.04, -0.01] (p = 0.004) but no significant change in weekly rates of hyperglycemia (−0.04 [95% CI: -0.10, 0.01]; p=0.102). The severe hypoglycemia events per 100 patients per year was 1.3 pre-implementation and 1.1 following implementation.

**Table 1.**
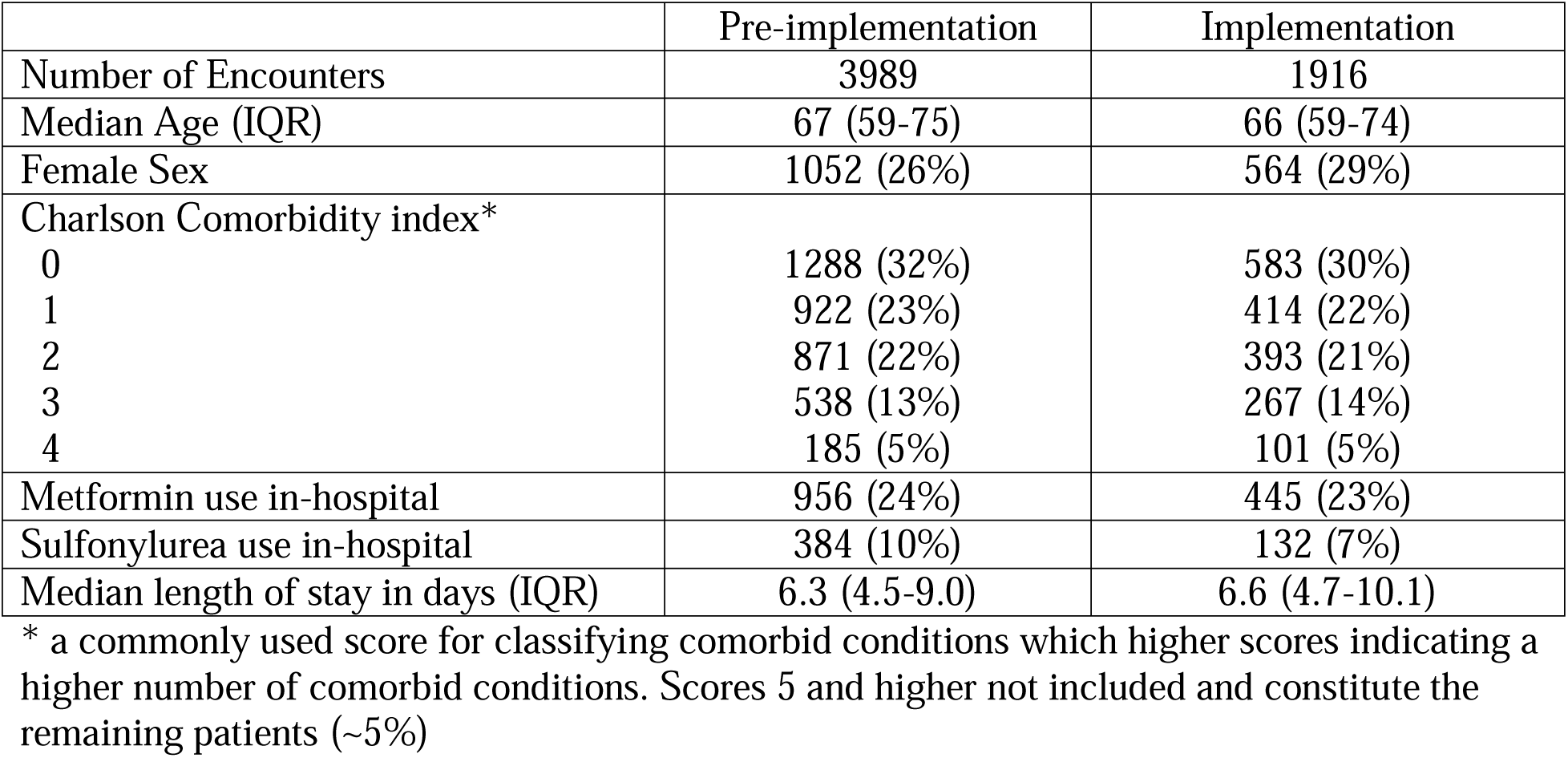
Baseline characteristics of included patients

**Figure 1.**
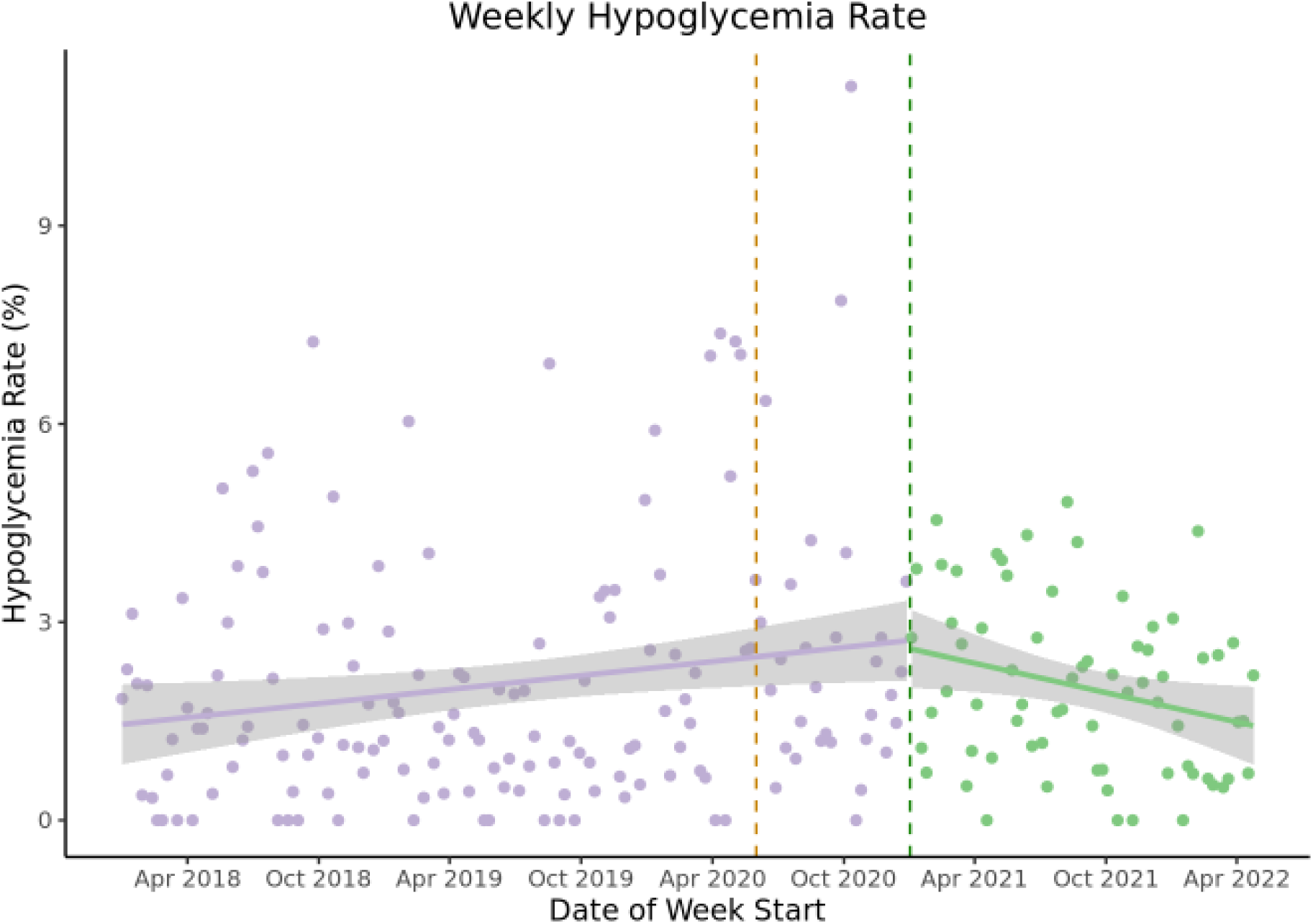
Rates of (A) hypoglycemia before and after model implementation Legend – the green dashed line indicates the time the multiple was fully implemented where as the dashed orange line indicates the start of the transition period.

## DISCUSSION

Our prospective analysis of a recently validated machine learned model^3^ to prevent hypoglycemia demonstrated a reduction in the rates of inpatient hypoglycemia. And while there are other studies that have sought to predict inpatient hypoglycemia,^6^ most have not been prospectively evaluated to assess their performance in routine care.

There are other strategies to prevent hypoglycemia in hospital such as having a virtual glucose management service.^7^ In a 1 year study including 3 hospitals in California, the implementation of this service, which consisted of a physician, nurse educator, and pharmacist, reduced rates of hypoglycemia by approximately 40% and the absolute number of severe hypoglycemic events (<2.2 mmol/L) was reduced from 47 per year to 15 per year following implementation (3.2 per 100 patient days to 1.0 per 100 patient days). The reduction in the rate of both hypoglycemia and severe hypoglycemia was impressive, but it is unclear how cost-effective, sustainable, or generalizable this model is. In contrast, our model does not require additional clinical team members and is constantly reviewing all of the available data for each patient, 24 hours per day and 7 days per week.

An important limitation of our study is that it occurred at a single hospital in Toronto, Ontario with only 1-year of data to evaluate its implementation. However, this is a necessary step before wider adoption to ensure the tool is achieving adequate performance. With only 1-year of implementation data we are likely under-powered to identify its impact on the rate of severe hypoglycemia because it is a rare event. Another limitation of all non-randomized studies is an inability to rule out unmeasured confounding or temporal changes that may have affected the primary outcome. For example, most of our implementation phase took place during the COVID-19 pandemic and prior data have shown that there was a marked reduction in the number of hospitalizations during this period for non-COVID related illness and increased severity of illness among those who did present with non-COVID related illness. Despite these limitations, the results of our study suggest that machine learning methods can be leveraged to prevent inpatient hypoglycemia.

## Data Availability

We cannot share the data

## Notes

**Conflicts of interest:** B.A.P. has received speaker honoraria from Abbott, Medtronic, Insulet, and Novo Nordisk; has served as an advisor to Insulet, Sanofi, and Abbott; and has received research support to his research institute from Novo Nordisk, and the Bank of Montreal (BMO). MF is a consultant for ProofDx, a start-up company that has created a point of care diagnostic tests for COVID-19 using CRISPR.

### Competing Interest Statement

B.A.P. has received speaker honoraria from Abbott, Medtronic, Insulet, and Novo Nordisk; has served as an advisor to Insulet, Sanofi, and Abbott; and has received research support to his research institute from Novo Nordisk, and the Bank of Montreal (BMO). MF is a consultant for ProofDx, a start-up company that has created a point of care diagnostic tests for COVID-19 using CRISPR

### Funding Statement

This study was funded by Banting and Best Diabetes Centre

### Author Declarations

The Unity Health Toronto Research Ethics Board gave ethical approval for this work (REB #16-371)

## References

1. Dayan I, Roth HR, Zhong A, et al. Federated learning for predicting clinical outcomes in patients with COVID-19. Nat Med. 2021;27(10):1735–1743.

2. Escobar GJ, Liu VX, Schuler A, Lawson B, Greene JD, Kipnis P. Automated Identification of Adults at Risk for In-Hospital Clinical Deterioration. N Engl J Med. 2020;383(20):1951–1960.

3. Fralick M, Dai D, Pou-Prom C, Verma AA, Mamdani M. Using machine learning to predict severe hypoglycaemia in hospital. Diabetes Obes Metab. 2021;23(10):2311–2319.

4. Diabetes Canada Clinical Practice Guidelines Expert Committee. Diabetes Canada 2018 Clinical Practice Guidelines for the Prevention and Management of Diabetes in Canada. Can J Diabetes. 2018;42 Suppl 1:S1–S5.

5. Levene H. Contributions to Probability and Statistics: Essays in Honor of Harold Hotelling. (Olkin I, ed.). Stanford University Press; 1960:278–292.

6. Mathioudakis NN, Abusamaan MS, Shakarchi AF, et al. Development and Validation of a Machine Learning Model to Predict Near-Term Risk of Iatrogenic Hypoglycemia in Hospitalized Patients. JAMA Netw Open. 2021;4(1):e2030913.

7. Rushakoff RJ, Sullivan MM, MacMaster HW, et al. Association Between a Virtual Glucose Management Service and Glycemic Control in Hospitalized Adult Patients: An Observational Study. Ann Intern Med. 2017;166(9):621–627.

